# A Sparse Gaussian Network Model for Prediction the Growth Trend of COVID-19 Overseas Import Case: When can Hong Kong Lift the International Traffic Blockade?

**DOI:** 10.1101/2020.05.13.20099978

**Authors:** Rui Miao, Qi Dang, Yong Liang

## Abstract

The COVID-19 virus was first discovered from China. It has been widely spread internationally. Currently, compare with the rising trend of the overall international epidemic situation, China’s domestic epidemic situation has been contained and shows a steady and upward trend. In this situation, overseas imports have become the main channel for china to increase the number of infected people. Therefore, how to track the spread channel of international epidemics and predict the growth of overseas case imports is become an open research question. This study proposes a Gaussian sparse network model based on lasso and uses Hong Kong as an example. To explore the COVID-19 virus from a network perspective and analyzes 75 consecutive days of COV-19 data in 188 countries and regions around the world. This article establishes an epidemic spread relationship network between Hong Kong and various countries and regions around the world and build a regression model based on network information to fit Hong Kong’s COV-19 epidemic growth data. The results show that the regression model based on the relationship network can better fit the existing cumulative number growth curve. After combining the SEIJR model, we predict the future development trend of cumulative cases in Hong Kong (without blocking international traffic). Based on the prediction results, we suggest that Hong Kong can lift the international traffic blockade from early to mid-June.

## Introduction

The COVID-19 virus was found in Wuhan, Hubei Province, China in December 2019. According to the evidence of early transmission dynamics, interpersonal communication has occurred between close contacts since mid-December 2019[1]. In order to control the spread of infections, Hubei and other provinces have adopted measures such as urban segregation and reducing inter-city mobility. Through a large number of public health interventions, the local epidemic situation in various provinces and cities in China has been basically controlled. However, the international spread of the epidemic is inevitable. Therefore, for areas where local transmission has been basically controlled, how to prevent overseas transmission has become the focus of current epidemic prevention work [2-4].

This article takes Hong Kong as an example to discuss how to effectively predict the cumulative case growth curve of regions with overseas imports as the main growth mode. As an important international transportation hub, the migration of a large number of international passengers has had an important impact on the spread of the epidemic in Hong Kong. Foreigners entering through international transportation channels such as aircraft shipping are the main way of increasing cases in the region. In order to reduce the possible transmission risk, from 0:00 on March 25, 2020, Hong Kong announced that non-Hong Kong people are prohibited from entering Hong Kong Airport. However, as an international financial and transportation center, Hong Kong will cause a lot of losses every day due to the international traffic blockade. Therefore, it is of great significance to predict the possible cumulative case growth rate of Hong Kong under the premise of incomplete blockade, and to further determine the possible date of unblocking in Hong Kong. However, the transmission rate in the traditional SIR/SEIR model is a constant value. However in practical problems, the transmission rate is constantly changing. For example, the growth rate of overseas imported cases is affected by changes in the international epidemic, etc. In this situation, it is difficult to use traditional infectious disease models to predict the future growth trend of Hong Kong[2,5]. Therefore, finding a new model to solve this problem has become an open research question.

In our research, we used 75 days of real-time infection data from 188 countries and regions around the world. Establish a case transmission relationship network between Hong Kong and other parts of the world through the sparse Gaussian network model based on lasso. The results show that the correlation coefficient between the epidemic trend in Hong Kong and several outbreak centers abroad is extremely high. At the same time, we can use the cumulative case growth data in areas with high correlation to Hong Kong in the network to establish a regression model to fit the cumulative case growth data in Hong Kong. After further combining the SEIJR model to predict case growth data of target areas (related to Hong Kong). We can predict the number of COVID-19 cases in Hong Kong without blocking traffic. Our findings can help Hong Kong adjust public interventions, estimate the time for lifting the blockade, and provide effective evidence to avoid serious outbreaks and economic losses.

## Materials and Methods

### Data source

The real epidemic data set used in this article mainly comes from the website:https://github.com/BlankerL/DXY-COVID-19-Data, Including the cumulative number of confirmed cases and cumulative number of cured cases from January 19 to April 2, 2020.

### Model

#### Sparse Gaussian network model

This article uses a Gaussian graph model based on Lasso to construct an international epidemic spread network. We use a Neighborhood selection strategy to solve the covariance selection problem. The specific model is as follows:

Consider n-dimensional multivariate normally distributed random variables *P =* (*P*_1_*,⋯⋯P_n_*) ~ *N*(*μ*,∑). This includes Gaussian linear models, for example,*P*_1_ is the response variable, {*P_k_*;2 ≤ *k* ≦ *n]* is a predictor. Assuming that the covariance matrix is nonsingular. We can use a graphical model *G =* (Γ,E) Conveniently express the conditional independence structure of the distribution, where Γ = {1,⋯⋯*n}* is the set of nodes, and E is the set of edges in P. Given all remaining variables *PΓ*\{*a*,*b*} = *{P_k_;k* ∈ Γ\{*a*,*b*}}, If and only if *P_a_* depends conditionally on *P_b_*, Only one pair (*a,b*) is included in the edge set E. Given all remaining variables, each pair of variables not included in the edge set is conditionally independent and corresponds to the zero term in the inverse covariance matrix [6].

Neighborhood selection is a subproblem of covariance selection. The minimum subset of the neighborhood Γ\{*a*}, of node *a∊Γ*, Therefore, considering all the variables in the neighborhood *P_a_, P_a_* conditionally independent of all remaining variables. The neighborhood of node *a∊*Γ consists of all nodes *b*∊Γ\{*a*}, Therefore (*a,b*)*∊*E. For the observation of, neighborhood selection aims to estimate (individually) the neighborhood of any given variable (or node). Neighborhood selection can be used as a standard regression problem. It can be effectively solved by Lasso [7], as shown in this article.

For sparse high-dimensional graphs, the consistency of the proposed neighborhood selection will be shown, the number of variables may increase with any power of the number of observations (high-dimensional), and the number of neighbors of any variable is the slowest than the number of observations (sparseness).

Neighborhood selection. As we all know, Lasso[7] proposed by Tibshirani et. Al., In the context of wavelet regression [8], it is called basic pursuit and has simplicity [8].When the forecast has all remaining variables *P_a_*{*P_k_*;*k*∊Γ\(*n*)\{*a*}}. The estimated value of the disappeared lasso coefficient asymptotically identifies the neighborhood of node a in the graph, as shown below. Let *n* × *p*_(_*_n_*_)_ dimensional matrix *p* contain *p* independent *n* observations. Therefore, for all *a∊*Γ(*n*), Column *P_a_* corresponds to a vector of *n* independent observations. Let〈▪,▪〉 be the usual inner product on Rn, and ‖▪‖_2_ is the corresponding norm [9].

Lasso estimates the formula of *θ^a^* in *θ^a,λ^* as formula (1):

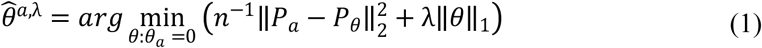

‖*θ*‖_1_ = ∑*b∊*Γ_(_*_n_*_)_|*θ_b_*| Is the *L*_1_ norm of the coefficient vector. It is recommended to normalize all variables to a common empirical variance in the above formula. The solution of the above formula is not necessarily unique. However, if the uniqueness fails, the solution set is still convex, and all of our results on the neighborhood apply to any solution of the above formula. Other regression estimates based on the *lp* norm have been proposed, where *p* is usually in the range [0,2] (reference[10]). A value of *p =* 2 will result in a ridge estimate and *p =* 0 corresponds to the traditional model selection. As we all know, only when *P* ≤ 1, the estimated value has a parsimony property (some components happen to be zero), For (*p*) ≥1, the optimization problem in the above formula is only convex. Therefore, the minimization of empirical risk constrained by *L*_1_ occupies *a* unique position, Due to *p =* 1 is the only value of *p*, the variable selection is performed on this value, and the optimization problem is still convex, so it is feasible for high-dimensional problems.

The neighborhood estimate (parameterized by *λ*) is defined by the nonzero coefficient estimate of *L*_1_ penalty regression as formula (2):

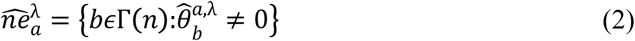

Therefore, each choice of penalty parameter *λ* specifies the estimate of the neighborhood of node *a*∊Γ(*n*), and the rest is to choose the appropriate penalty parameter. A larger penalty value tends to reduce the size of the estimated set, and if the value of *λ* decreases, usually more variables are included in the estimated value.

Predict Oracle solutions. A seemingly useful choice of penalty parameters is (unavailable) to predict the oracle value as formula (3):

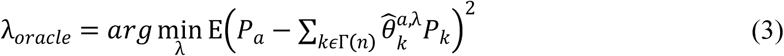

Expectation is understood to be about the new *P*, which has nothing to do with the samples that estimate *θ^a^*^,λ^. The prediction penalty minimizes the prediction risk in all Lasso estimates. The λ*oracle* estimate is obtained by selecting λ*cv* for cross-validation.

Shao[11] showed that for a 10-penalty return. The cross-validation selection of penalty parameters is consistent with the model selection of the verification set size under certain conditions. Predict that the Oracle solution will not lead to consistent model selection for Lasso. Proposition 1. Let the number of variables grow to infinity. For *n → ∞,p*(*n*) *→ ∞*, and *γ >* 0,*p*(*n*) = *o*(*n^γ^*). Suppose the covariance matrix ∑(*n*) except for some pairs(*a*,*b*)∊Γ(*n*) × Γ(*n*). In ∑*_ab_*(*n*) = ∑*_ba_*(*n*) = *s*. For some 0 < *s* < 1 and all *n∊N*. Under the prediction oracle penalty, the probability of choosing the wrong neighborhood for node *a* converges to 1 as formula (4):

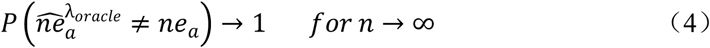

From the proof of Proposition 1, it can be concluded that many noise variables are included in the prediction of the neighborhood of the Oracle solution. In fact, for a fixed number of variables, the possibility of including noise variables in prediction predictions will not even disappear gradually. If the selected penalty is greater than the predicted optimal value, then Lasso can be used for consistent neighborhood selection.

#### Regression model

This article uses Passive Aggressive Algorithms as the regression model for this study. Passive attack algorithms are a class of algorithms for large-scale learning. Similar to the perceptron, it does not need to set the learning rate. However, there is an additional regularization parameter C than the perceptron. Similar to most regression algorithms, the purpose of Passive Aggressive Algorithms is to use training samples to learn relevant parameters to minimize the value of the loss function [12]. In the learning of training samples (*x*,*y*) one by one, Passive Aggressive Algorithms uses stochastic gradient algorithms to update parameters. First, the gradient ∇*J* of the loss J associated with the newly input training samples (*x*,*y*) is obtained. Then update the parameter *θ* in the direction of gradient descent as formula (5):

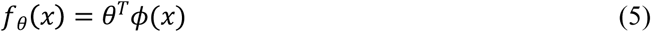

In the probability gradient descent method, When the gradient descent is too large, the learning results tend to be unstable; When the gradient descent is too small, it will make the convergence rate slower.

If researchers can reasonably choose the loss function. It can make the gradient drop to the bottom of the valley quickly. Therefore, a penalty coefficient is generally introduced. That is, when deviating from the current solution *θ*, make appropriate adjustments to the amount of gradient descent. Therefore, we can get formula (6):

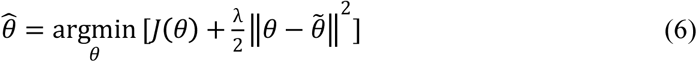

*λ* is a positive scalar. Such a learning method can effectively suppress the gradient descent. This Algorithms is called Passive Aggressive Algorithms.

The specific algorithms of Passive Aggressive Algorithms are:

1. Select the initial value, *θ* ← 0
2. Using the newly input training samples (x, y), formula 6 is updated for the parameter *θ* as formula (7).

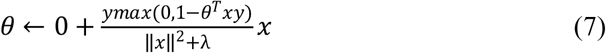
3. Repeat the second step until convergence In the regression problem, the function J (0) has two different ways: using *L*_1_ − *norm* and *L*_2_ − *norm*. As shown in formula (8-9):

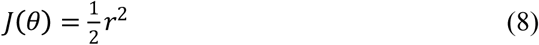

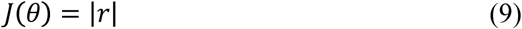 This article uses two regularizes to establish regression models respectively and uses data from countries and regions related to Hong Kong to fit Hong Kong’s COV-19 cumulative cases data.

#### SEIJR model

After establishing regression models using known data. We found that most areas related to Hong Kong are still in the outbreak period of local transmission. In this situation, we can use the traditional infectious disease model to predict the growth trend of cases in these areas, thereby further predicting the possible growth trend of Hong Kong (without international traffic blockade). Therefore, this article first uses the SEIJR model to predict the growth curve of the number of local diagnoses in countries and regions related to Hong Kong. Finally, through these data, we can fit and predict the future growth trend of Hong Kong.

The classic SEIR model divides the crowd into S (Susceptible), I (Infected), E (Exposed) and R (Recovered)[13]. The model also assumes that all individuals in the population have an organic infection rate[14,15]. When the infected individual recovers, antibodies will be produced, that is, the recovered population R will not be infected again.

In this study, the population was classified as susceptible S, latent E, infectious I, diagnosed J, recovered R, *S*_1_ and *S*_2_ respectively representing two populations with different susceptibility among susceptible populations, The infection risk of *S*_2_ is low, risk probability value is p, the probability of asymptomatic latent persons being infectious is q, the probability of latent persons turning into infected persons is k, the isolation rate is l, the confirmed rate of infectious persons is α, the infectious person’s The recovery rate γ1, the fatality rate of the infectious person δ, the recovery rate of the diagnosed person γ2, the mortality rate δ[16]. The transmission rate β is defined as the average number of infections caused by a person who is susceptible to contact with Class I per unit time. Based on the above parameters, the model is established as follows as formula (10-15):

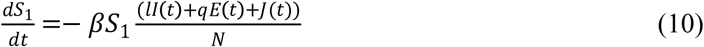

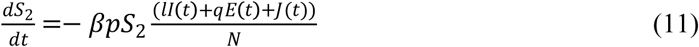

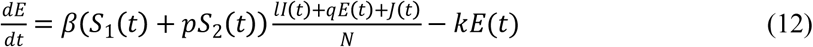

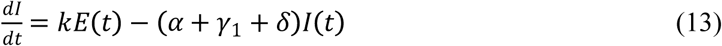

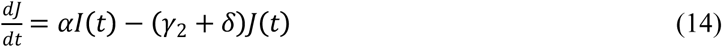

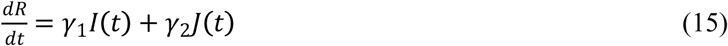

## Results

In this chapter, we first use the Gaussian sparse network model based on lasso to build a global relationship network. Then, extract the Hong Kong subnet. Next, use the Passive Aggressive Algorithm to establish a regression model to fit the existing cumulative case data. The experment use 5-flod cross-validation, explained_variance, mean_absolute_error and r2 Four indicators to verify the model results. Finally, the SEJIR model is used to predict the growth trend of the number of people in areas related to Hong Kong and to predict the cumulative case growth curve of Hong Kong without blocking international traffic.

### Establish an epidemic relationship network

As shown in Figure 1, this article first establishes a relationship network for the spread of epidemics in 188 countries and regions around the world, because the Hong Kong government promulgated international traffic blockade measures on March 25, Beijing time. Announced the ban on the entry of international tourists other than Hong Kong nationals. Therefore, considering that the COV-19 virus has the longest incubation period of 14 days. This article takes 7 days from March 25 onwards, which is April 2 as a time node for establishing an international relationship network. Establish an international network of 75 days from January 19 to April 2.

**Figure 1.**
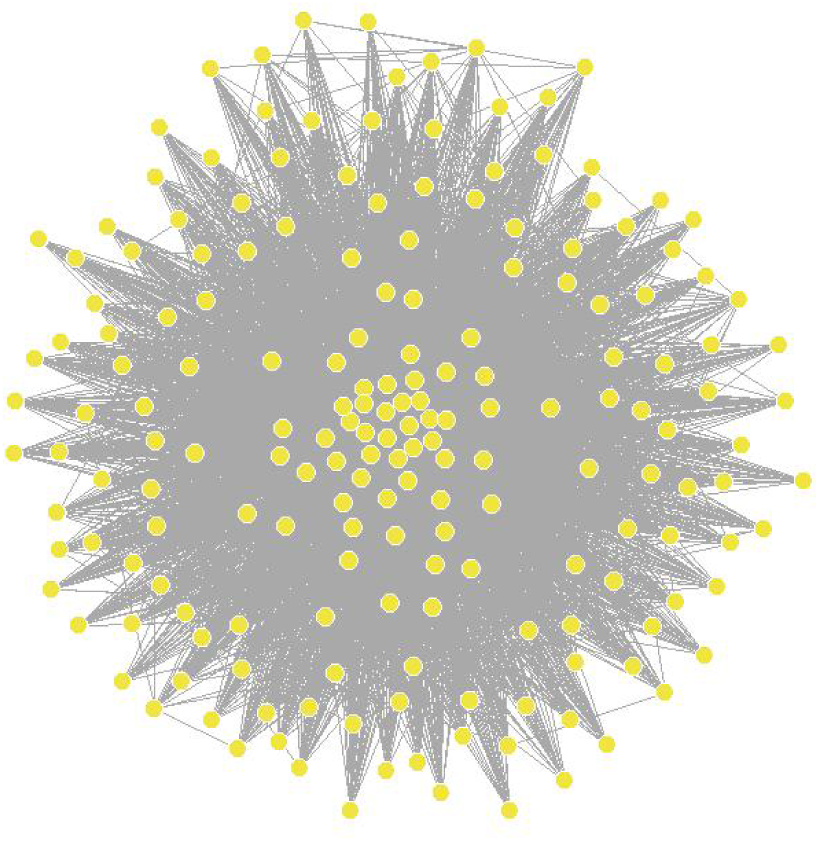
International epidemic relations network in 188 countries and regions

The results are encouraging, From Table 1, we find that the United States, Hubei, China, Iran, Italy and other countries are Hub nodes in the network. These countries have the most relations with other countries, which means that these countries are the main sources of international communication. This result is consistent with existing general knowledge.

**Table 1.** Hub nodes in the international epidemic relationship network

|  |  |  |  |  |  |  |  |
| --- | --- | --- | --- | --- | --- | --- | --- |
| Country | United States | Hubei, China | Iran | Italy | Germany | France | Spain |
| Number of nodes | 186 | 186 | 186 | 186 | 184 | 184 | 182 |

Next, we extracted the subnets of countries and regions related to Hong Kong. As shown in Figure 2, we found that 24 countries and regions are connected to Hong Kong. In addition to the hub nodes of the network. We also found South Korea, Russia and other countries that have close contacts with Hong Kong. We hope that the use of data from these countries and regions that are closely related to Hong Kong can help us fit the existing cumulative case growth data in Hong Kong and predict future trends. We have found that in areas related to Hong Kong, except for a few areas such as Hubei in China, other areas are at the peak of local transmission. This situation means that researchers can use traditional infectious disease models to predict epidemic trends in these areas, which is also the basis for this article to use the SEJIR model.

**Figure 2.**
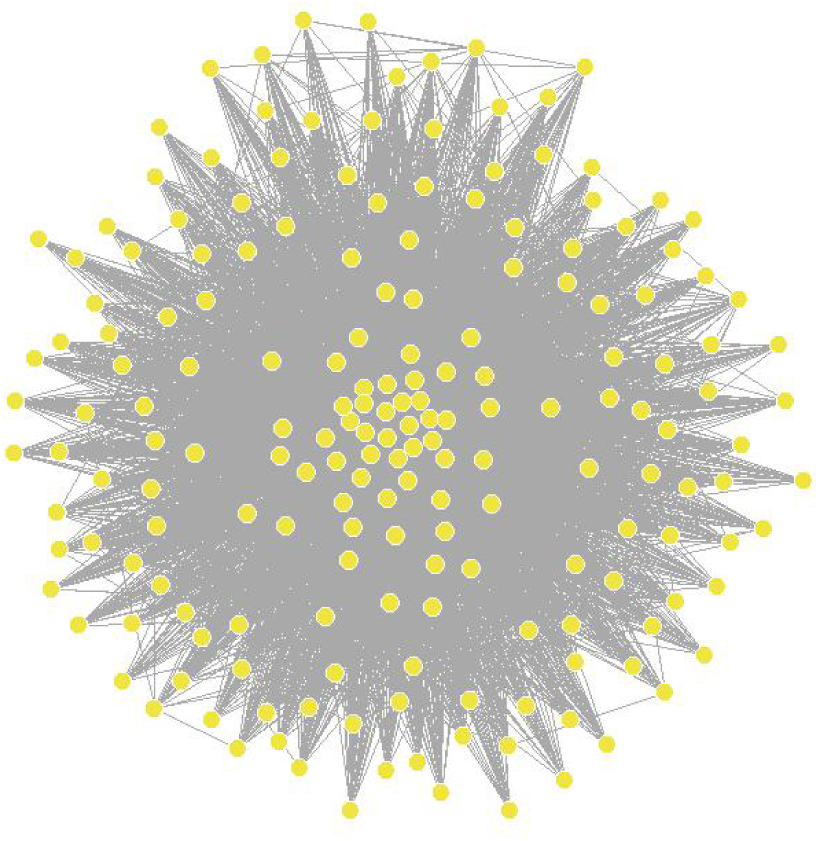
International epidemic relations networks in 24 countries and regions related to Hong Kong

### Fit existing case growth

After getting the Hong Kong relationship subnet. We used Passive Aggressive Algorithm based on *L*_1_ and *L*_2_ norm to establish regression models respectively. The experimental results are shown in Figure 3. As shown in Table 2-3, the Passive Aggressive Algorithm can fit the existing growth curve well, and the error of the 5-fold cross-validation is only -6.94. Among them, Passive Aggressive Algorithm_L2 has a better realization, and the minimum error is only 0. 12. The explained_variance and r2 indicators of both models are 0.99, and mean_absolute_error is lower than 13.3, Shows an extremely high degree of fit to existing growth data. This result shows that we can use the data based on the relation network to fit the existing case growth situation in Hong Kong. This means that we can use 24 countries related to Hong Kong to predict the future case growth curve of Hong Kong (unblocked traffic).

**Figure 3.**
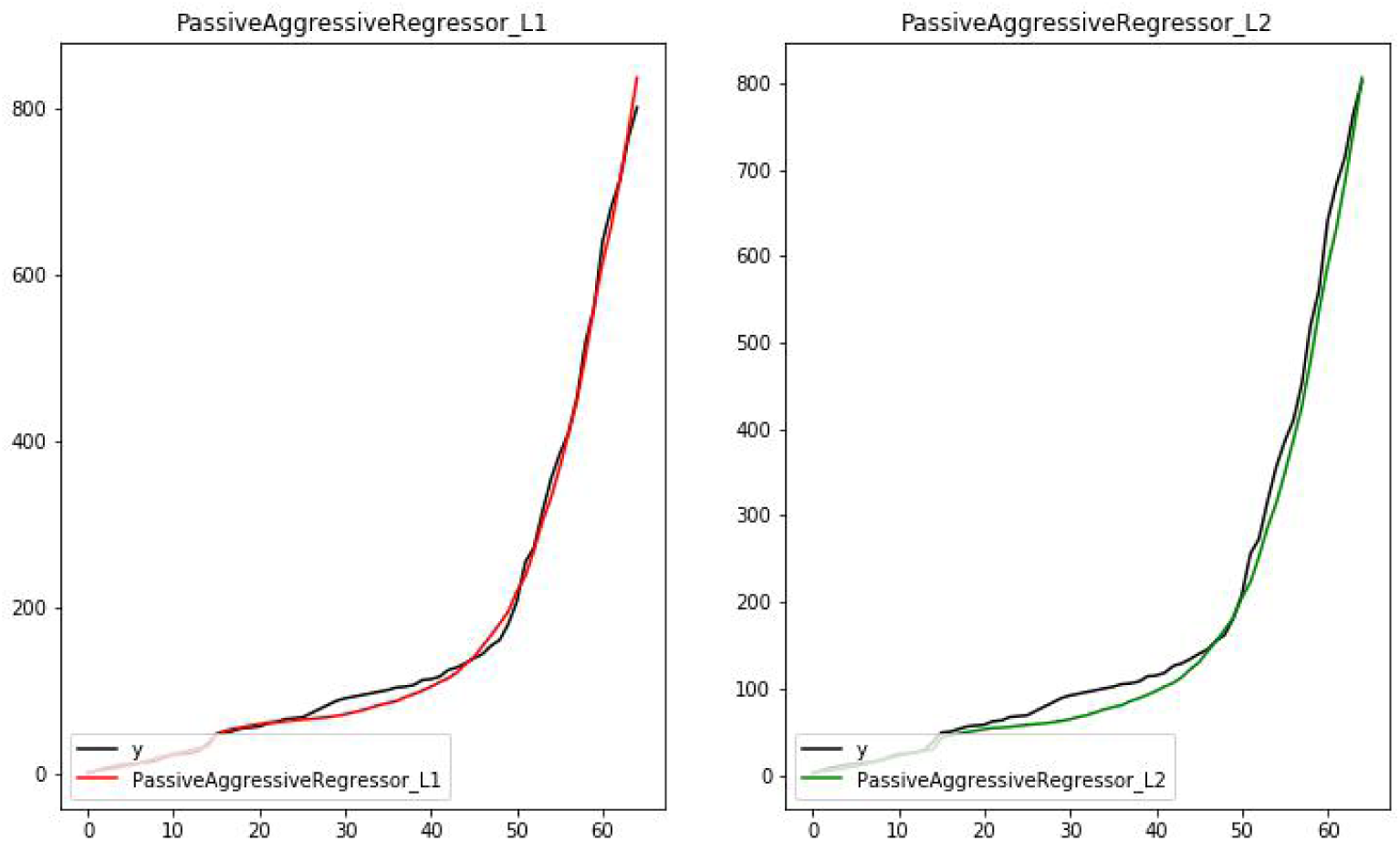
Fitting curve using Passive Aggressive Algorithm based on *L*_1_ and *L*_2_

**Table 2.** Passive Aggressive Algorithm *L*_1_ and *L*_2_ based on 5-fold cross-validation results

|  | 0 | 1 | 2 | 3 | 4 |
| --- | --- | --- | --- | --- | --- |
| **PassiveAggressiveRegressor_L1** | 0.97 | -1.06 | -6.94 | 0.6 | 0.6 |
| **PassiveAggressiveRegressor_L2** | 0.12 | -1.40 | -0.47 | 0.63 | 0.55 |

**Table 3.**
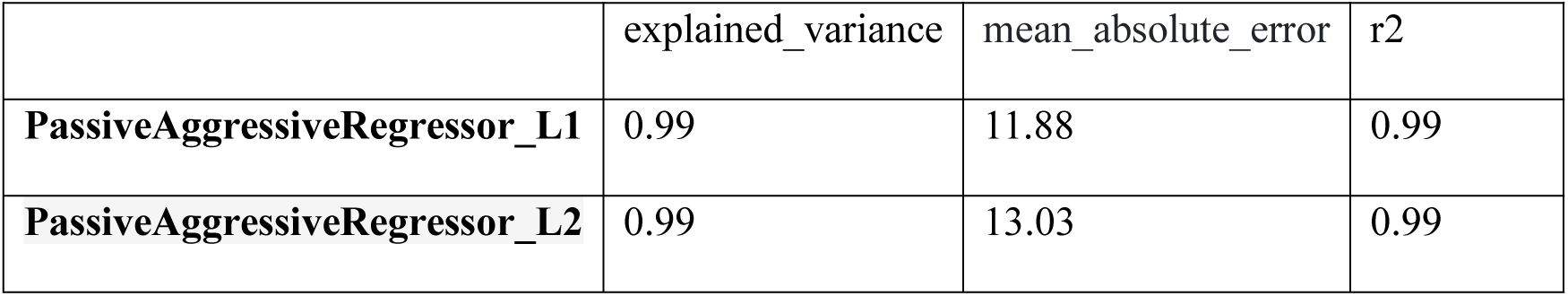
Passive Aggressive Algorithm *L*_1_ and *L*_2_ performance of different indicators

### Predict the growth of cases in Hong Kong (unblocked traffic)

Finally, we used the SEJIR model to simulate the cumulative case growth data of 23 countries and regions related to Hong Kong. The parameters used in this article are shown in Table 4, where Beta is a floating value, which is adjusted according to the specific conditions of each country. Since this article can collect real data from other countries from April 2-19, this article first uses real data to predict the growth curve of Hong Kong’s epidemic without blocking international traffic, the purpose is to evaluate the necessity of traffic blockade. As shown in Figure 4, if international traffic is not blocked, the model shows that as of April 19, the cumulative number of cases in Hong Kong is likely to be close to 1,800. However, after the traffic blockade, there are only about 1,000 cases in Hong Kong. This shows that under the current circumstances, the blockade of Hong Kong is necessary.

**Table 4.**
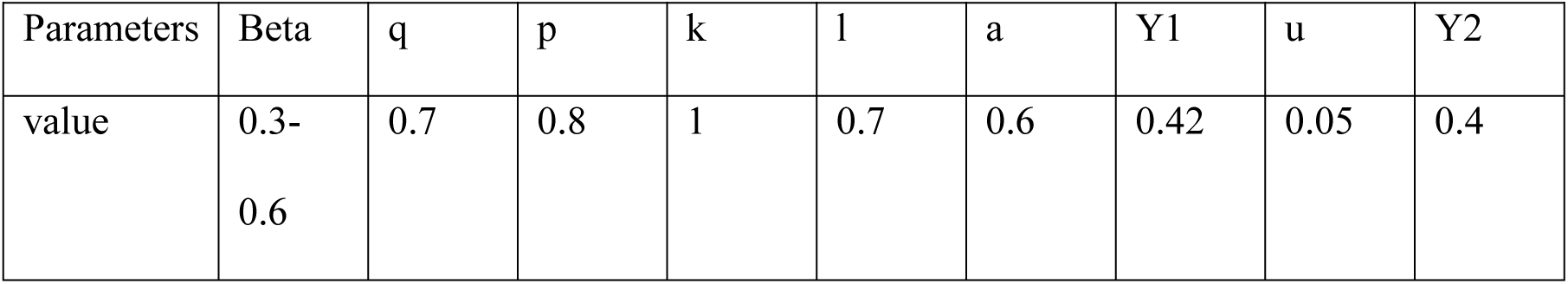
Parameters of SEJIR model

|  |  |  |  |  |  |  |  |  |  |
| --- | --- | --- | --- | --- | --- | --- | --- | --- | --- |
| Parameters | Beta | q | p | k | l | a | Y1 | u | Y2 |
| value | 0.3-0.6 | 0.7 | 0.8 | 1 | 0.7 | 0.6 | 0.42 | 0.05 | 0.4 |

**Figure 4.**
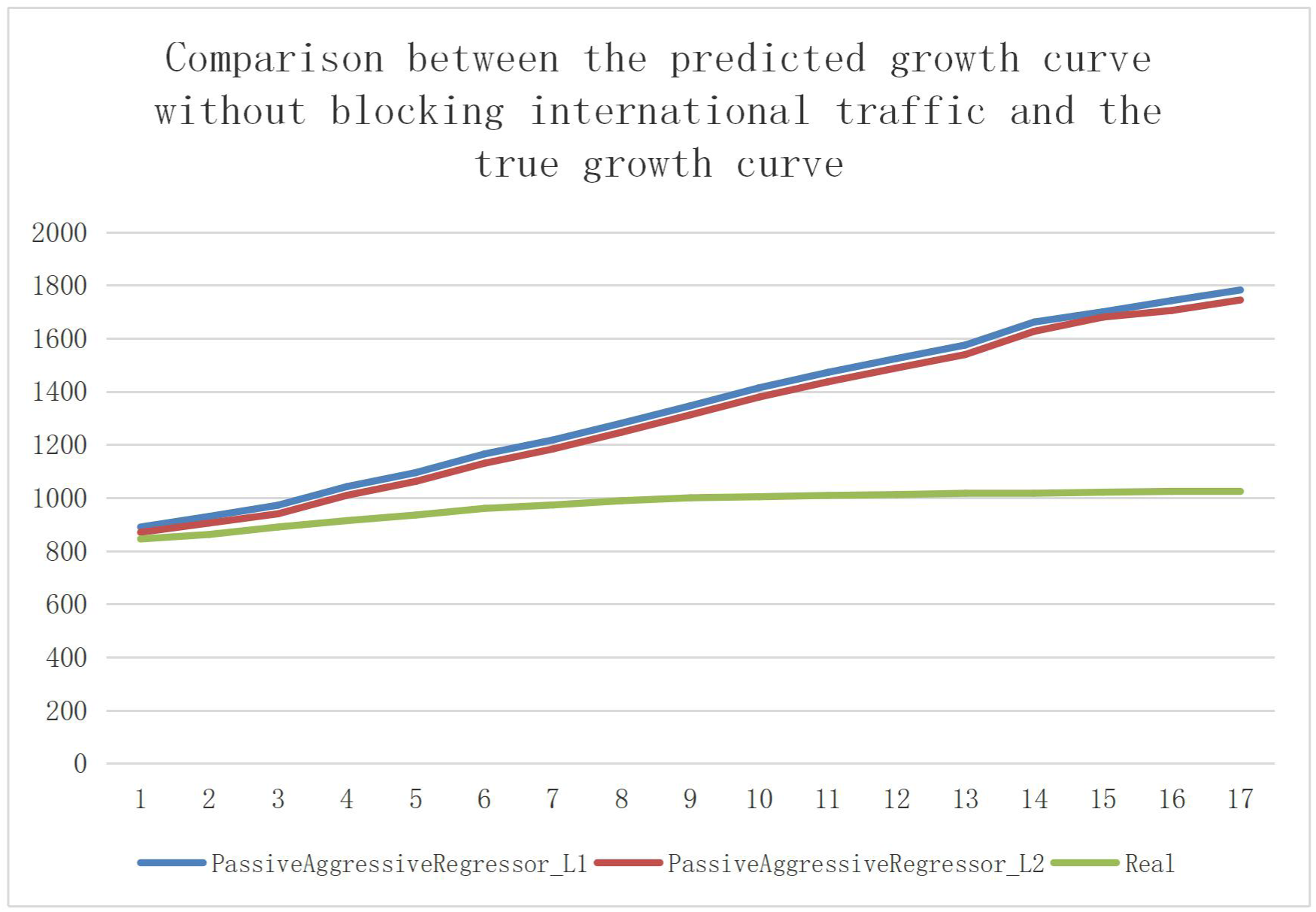
Comparison between the predicted growth curve and the true growth curve without hindering international flows

Figure 5 is the model’s forecast of cumulative case growth in Hong Kong (unblocked traffic), and Figure 6 is the daily new case curve. The model shows that if traffic is not blocked, the cumulative cases in Hong Kong may eventually exceed 7,000, and the rapid growth of cumulative cases will continue for more than 30 days. New cases in a single day reach a peak in late April, and then gradually decline. However, after 40 days, new cases may be reduced to single digits in a single day. Based on the model results, we suggest that the date of lifting the blockade in Hong Kong should not be earlier than the end of May. Considering the incubation period of the virus and the risk of asymptomatic, the best unsealing date should be until early to mid-June.

**Figure 5.**
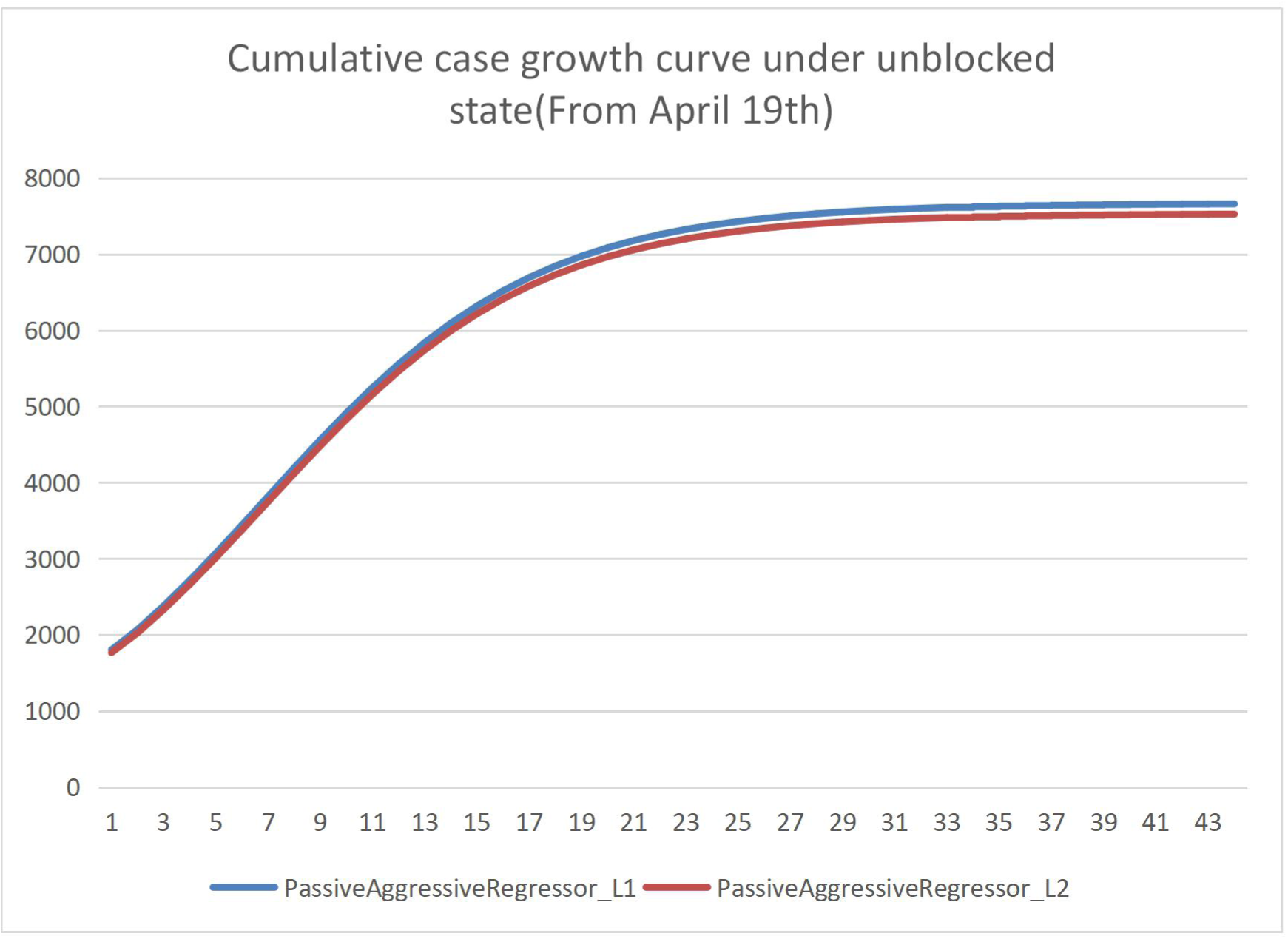
Cumulative case growth curve under smooth conditions (since April 19)

**Figure 6.**
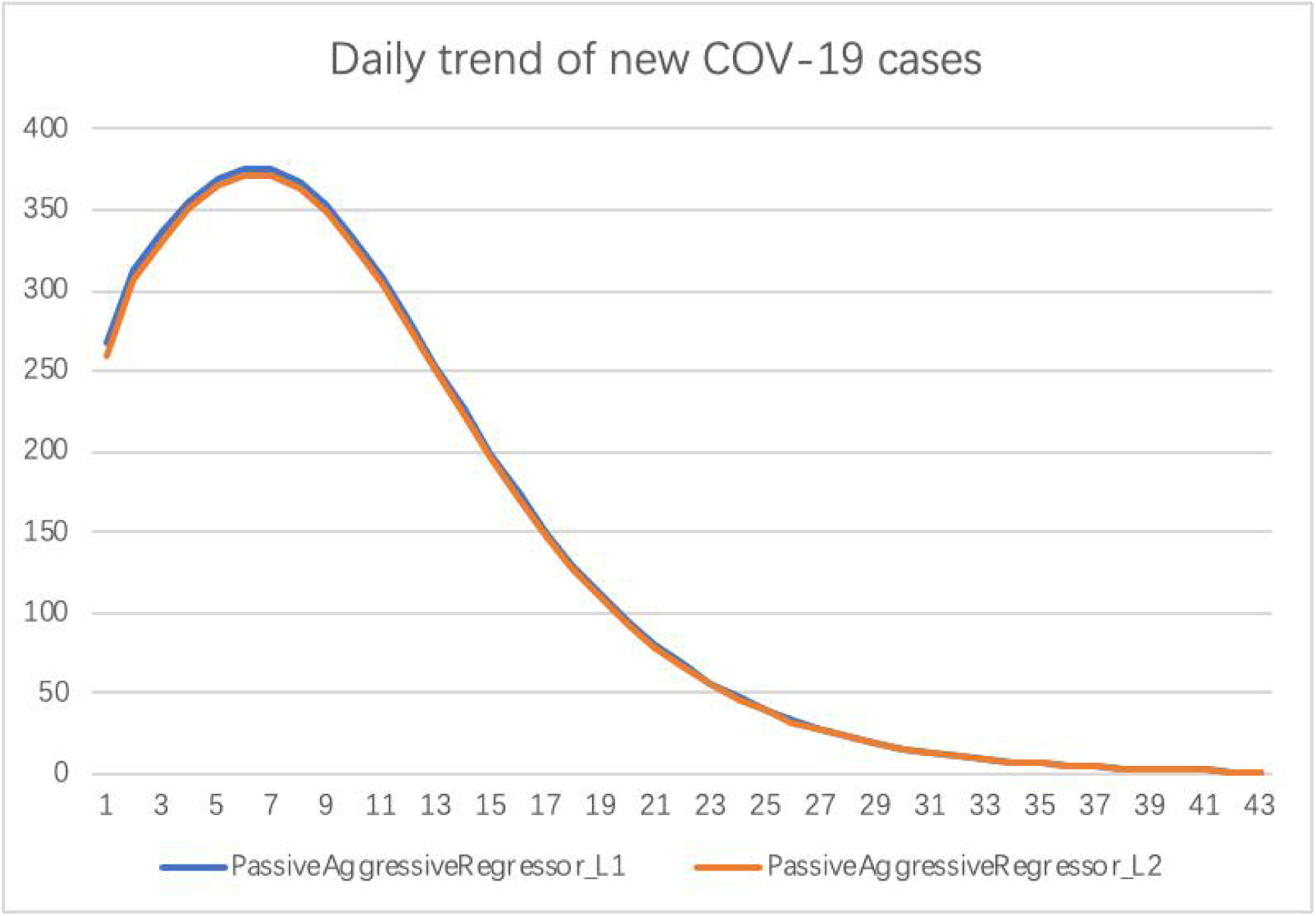
COVID-19 daily growth trend

## Discussion

At present, researchers have done a lot of work to predict the trend of local transmission of COV-19. However, in many countries and regions, the growth pattern of outbreaks is dominated by overseas imports. Existing research is difficult to predict effectively. Therefore, we take Hong Kong as an example, the purpose is to propose an infectious disease model that predicts the growth trend of imported cases abroad.

In this article, first, we proposed the sparse network model based on lasso, by analyzing the data matrix of real case statistics and drawing the COV-19 epidemic international network. From this network, we can get the countries related to the Hong Kong epidemic. Second, we introduce the Passive Aggressive Algorithm and SEJIR model. After obtaining the Hong Kong subnet information, we use the Passive Aggressive Algorithm to establish a regression model and use the data of countries and regions related to Hong Kong in the relationship network to fit the existing growth data of Hong Kong. According existing COV-19 case data and literature. We can determine the parameters of the model, such as recovery rate, mortality rate, diagnosis rate, etc., We use SEJIR model to predict the epidemic growth trend in areas relevant to Hong Kong, such as United States, Italy, etc. Based on the results of the SEJIR model, we can finally use the Passive Aggressive Algorithm to predict the future development trend of Hong Kong. We hope this article can help the Hong Kong government reduce the losses caused by the COV-19 epidemic, and also provide an international epidemic spread prediction model for areas similar to Hong Kong.

This article still has some shortcomings. For example, we assume that international tourists in Hong Kong will be quarantined and all infected persons will be diagnosed. However, some asymptomatic infected persons may still be missed, causing local transmission in Hong Kong. Therefore, in the next step we plan to count the asymptomatic infection rate in Hong Kong and further improve the model in this article.

## Data Availability

The real epidemic data set used in this article mainly comes from the website:https://github.com/BlankerL/DXY-COVID-19-Data，Including the cumulative number of confirmed cases and cumulative number of cured cases from January 19 to April 2, 2020

## Declaration of intesrst statement

No potential conflict of interest was reported by the author(s).

